# Artificial Intelligence Predicts and Explains West Nile Virus Risks Across Europe: Extraordinary Outbreaks Determined by Climate and Local Factors

**DOI:** 10.1101/2020.07.24.20146829

**Authors:** Albert A Gayle

**Affiliations:** Department of Public Health and Clinical Medicine, Section of Sustainable Health, Umeå University, SE-90187 Umeå, Sweden

## Abstract

Year-to-year emergence of West Nile virus has been sporadic and notoriously hard to predict. In Europe, 2018 saw a dramatic increase in the number of cases and locations affected. In this work, we demonstrate a novel method for predicting outbreaks and understanding what drives them. This method creates a simple model for each region that directly explains how each variable affects risk. Behind the scenes, each local explanation model is produced by a state-of-the-art AI engine. This engine unpacks and restructures output from an XGBoost machine learning ensemble. XGBoost, well-known for its predictive accuracy, has always been considered a “black box” system. Not any more. With only minimal data curation and no “tuning”, our model predicted where the 2018 outbreak would occur with an AUC of 97%. This model was trained using data from 2010-2016 that reflected many domains of knowledge. Climate, sociodemographic, economic, and biodiversity data were all included. Our model furthermore explained the specific drivers of the 2018 outbreak for each affected region. These effect predictions were found to be consistent with the research literature in terms of priority, direction, magnitude, and size of effect. Aggregation and statistical analysis of local effects revealed strong cross-scale interactions. From this, we concluded that the 2018 outbreak was driven by large-scale climatic anomalies enhancing the local effect of mosquito vectors. We also identified substantial areas across Europe at risk for sudden outbreak, similar to that experienced in 2018. Taken as a whole, these findings highlight the role of climate in the emergence and transmission of West Nile virus. Furthermore, they demonstrate the crucial role that the emerging “eXplainable AI” (XAI) paradigm will have in predicting and controlling disease.

**Highlights:**

- This study shows that the extraordinary 2018 West Nile virus outbreak in Europe was likely due to cross-scale effects between large climatic systems and local mosquito vector populations
- We found that large areas in Europe are similarly vulnerable to large and sudden outbreaks
- These findings were powered by a novel AI-driven engine for deriving locally precise models; this explanatory engine was supported by a high-performance XGBoost model (97% AUC).
- AI-driven local models allow for high-power statistical analyses, including: hypothesis testing,, standardized effect size calculation, multivariate clustering, and tertiary inferential modeling

## Background

### Predicting West Nile virus in context

West Nile virus (WNV) is a versatile pathogen that is amplified in the environment via spread among local animal populations, most notably birds. Mosquitoes serve as the primary vector and facilitate transmission between reservoir hosts and on to humans. Many other potential vectors, amplification hosts, and zoonotic transmission routes have been implicated^1,2^. Infected humans also directly contribute to disease spread, via several confirmed routes^3^. In Europe, 2010 seemed to signal a new epidemiological phase: a new more virulent form of the previously mild lineage 2 variant emerged, supplanting the original in terms of morbidty and mortality, and geospatial spread^4^. And this expansive trend has continued into 2018 and beyond^5^. 2018 saw a 7.2 fold increase in reported cases and a markedly expanded geographic range (see *Figure 1*). 2019 then saw the first autochthonous human cases in Germany leading to concerns of sudden and accelerating spread^6^. Even mild manifestations of infection are liable to be misdiagnosed or missed^7^, resulting in wide-scale underreporting^8^. Suggestions of potential late onset functional and/or cognitive deficits among the seemingly healthy^9^, have furthermore fueled concerns. Recent seroprevalence studies suggest human infection rates between 1-3%, and as high as 6%, in in parts Europe^10-12^. And with seropositivity as high as 90% in endemic sub-Saharan Africa^13^, the potential for expansion is very real.

The spread of WNV among humans depends on a wide range of determinants, including climatic features, environment, and sociodemographic factors^14^. However, efforts to quantify these relationships are often confounded by complex interactions and time-varying associations^15^. To resolve such issues, some have suggested higher-dimensional models effected at local scales^16-18^.

In this paper, we demonstrate one such solution. Our solution is powered by a novel Al-based engine, the SHAP (SHaply Additive Explanation) framework^19^. SHAP uses the outputs of a popular classification tree ensemble, XGBoost^21^, to generate local explanatory models for each individual case^20^. These models are statistically robust. They furthermore possess many favorable properties that allow for robust, hypothesis-driven summarization and analysis. Our aim was therefore to evaluate this solution in the context of geospatial modeling and prediction of infectious disease. To this end, the extraordinary WNV outbreak of 2018 in Europe was selected as an ideal test case.

## Results

The 2018 WNV outbreak season was extraordinary in many ways. The number of regions reporting cases as well as the proportion of regions affected per country was substantially higher than in previous years *(Figure 1a)*. Beyond that, many regions affected in 2018 had a history of WNV outbreak, but emergence has been overall sporadic *(Figure 1b)*. Overall, observed WNV range was substantially higher in 2018 compared to control (134 regions vs 44.9 mean regions during the 2010-2016 training period; see *Figure 1c)*, with 25.4% previously naive to WNV.

**Figure 1.**
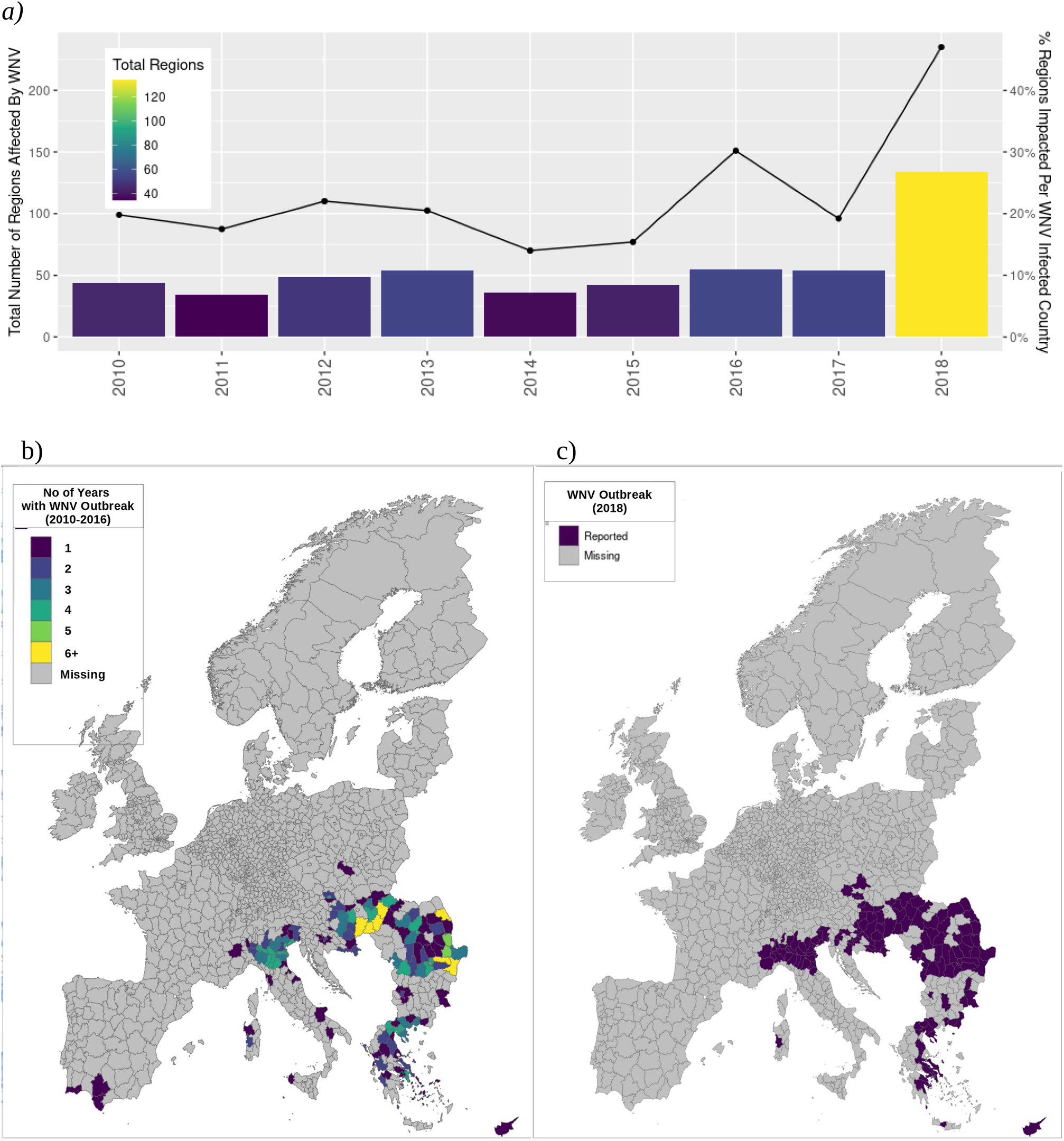
Spatiotemporal distribution of WNV infection and ecoclimatic trends. Our study included 1811 NUTS3 subregions comprising 29 countries within the EU/EEA (*Supplementary Table 1*). During the study period, WNV outbreaks were found to occur sporadically but with an overall increasing trend in terms of number of WNV cases reported per affected country per year and number of regions reporting WNV cases per affected country per year. This trend was driven primarily by the extraordinary 2018 outbreak year in which more cases were reported than in the preceding 7 years combined. *(Figure 1a)*. The geospatial spread of WNV infection during the 2018 event year (*Figure 1b)* is broadly equivalent to the cumulative range from the 2010-2016 control period *(Figure 1c)*.

### Out-of-sample predictive results were found to be exceptional

Our XGBoost model delivered an AUC of 0.97. This is particularly notable given the substantial proportion of previously naive regions affected in 2018 and overall sporadic nature of occurrence during the training period (2010-2016). Refer to *Figure 1*. Sensitivity of the threshold-optimized model was .89 and balanced accuracy was .92, which is noteworthy given the overall sparsity of positive outbreak regions.

**Figure 2.**
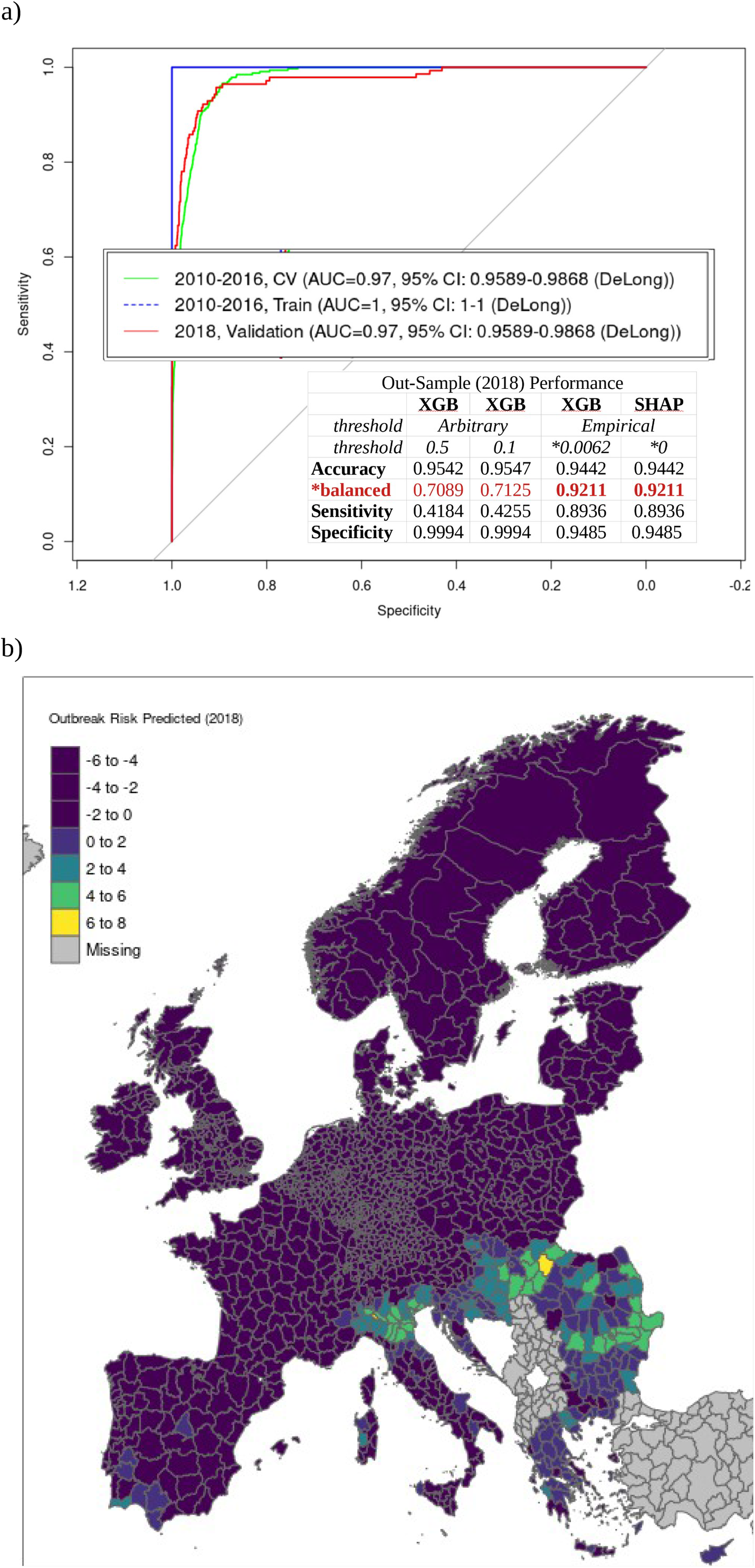
Local model estimation enhances predictive power. A generic model for predicting WNV outbreak range in Europe was generated using XGBoost. Refer to *Methods* for details. Cross-validation was initially used to determine the optimal number of recursive trees at which the holdout deviance was minimized. This iteration value (872) was doubled and applied to an initial training run. Of the original n=1332 original features, only n=427 were determined to be relevant. This figure included n=100 (from an initial n=1811) geospatial covariates included for control. Another model was then trained with the selected (n=427) feature set. The extraordinary 2018 outbreak was used for out-of-sample validation. AUC performance was found to be extremely high (1.00 and 0.97 in-sample and out-sample, resp.) – on par with diagnostic cross-training results *(Figure 2a)*. SHAP was then applied to predict feature effects for each region. The product of this reanalysis is a system of additive equations dimensionally identical to the original data set. Each feature effect term reflects its independent contribution to the local log relative risk of outbreak, conditioned on the global baseline risk. This global baseline is calculated automatically by SHAP and also serves to recenter the estimations. This "BIAS" is the error term inherent to methods of linear approximation and indicates the point at which the function crosses the axis; and therefore makes for an empirically ideal discriminatory threshold. This was calculated to be -5.0822, which implies a baseline probability of 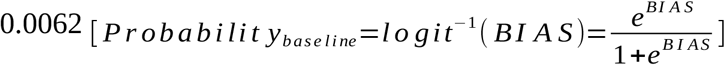. Summing each local SHAP model therefore produces an estimate of the degree to which outbreak is indicated or contraindicated (log relative risk), for a given region. Positive discriminatory performance for the optimized SHAP model was substantially improved, with sensitivity more than twice that of the XGBoost models with standard threshold values (*Figure 2a*). The SHAP model outputs were confirmed to be identical to the XGBoost outputs when the SHAP-estimated baseline probability was used as the discriminatory threshold. That said, the SHAP model provides uniformly scaled, bipolar output – which allows for clearer interpretation and enhanced geospatial analyses (*Figure 2b*).

### Observed feature effects dependent on geospatial scale

The nature of ensemble models precludes direct assessment of feature effects. XGBoost is no different. Feature importance can however be assessed indirectly. Refer to *Methods* and *Figure 3*. “Gain” is the amount by which global predictive power decreases upon removal of a given feature from the model. The feature describing year-to-year correlation, “Recent History of WNV [past year]”, was found to contribute the most by this metric (6.3%). This was followed closely by “maximum temperature of the warmest month” (5.6%). No other features achieved similar levels of imputed global importance. Substantial differences were observed between gain and two other importance criteria, “frequency” and “coverage”. For example, our largest contributor to predictive output, year-to-year autocorrelation, was found to impact only a small minority of ensemble outcomes (low “frequency”). And the features found to be most consistently relevant case to case (high “coverage”) were only moderately impactful in terms of gain. Considered simultaneously, few features rank highly in terms of all three criteria (Figure 3, first quadrant) – maximum temperature of the warmest month, vapor pressure in the second quarter and maximum temperature in the third quarter – all climatic. Features associated with hosts, vectors, and spatial covariates were found to be relevant only with respect to a limited set of regions (low coverage and frequency; third quadrant of *Figure 3*). Sociodemographic, environmental, and economic features perform marginally better in terms of coverage, indicating effect spanning multiple regions but far from globally consistent.

**Figure 3.**
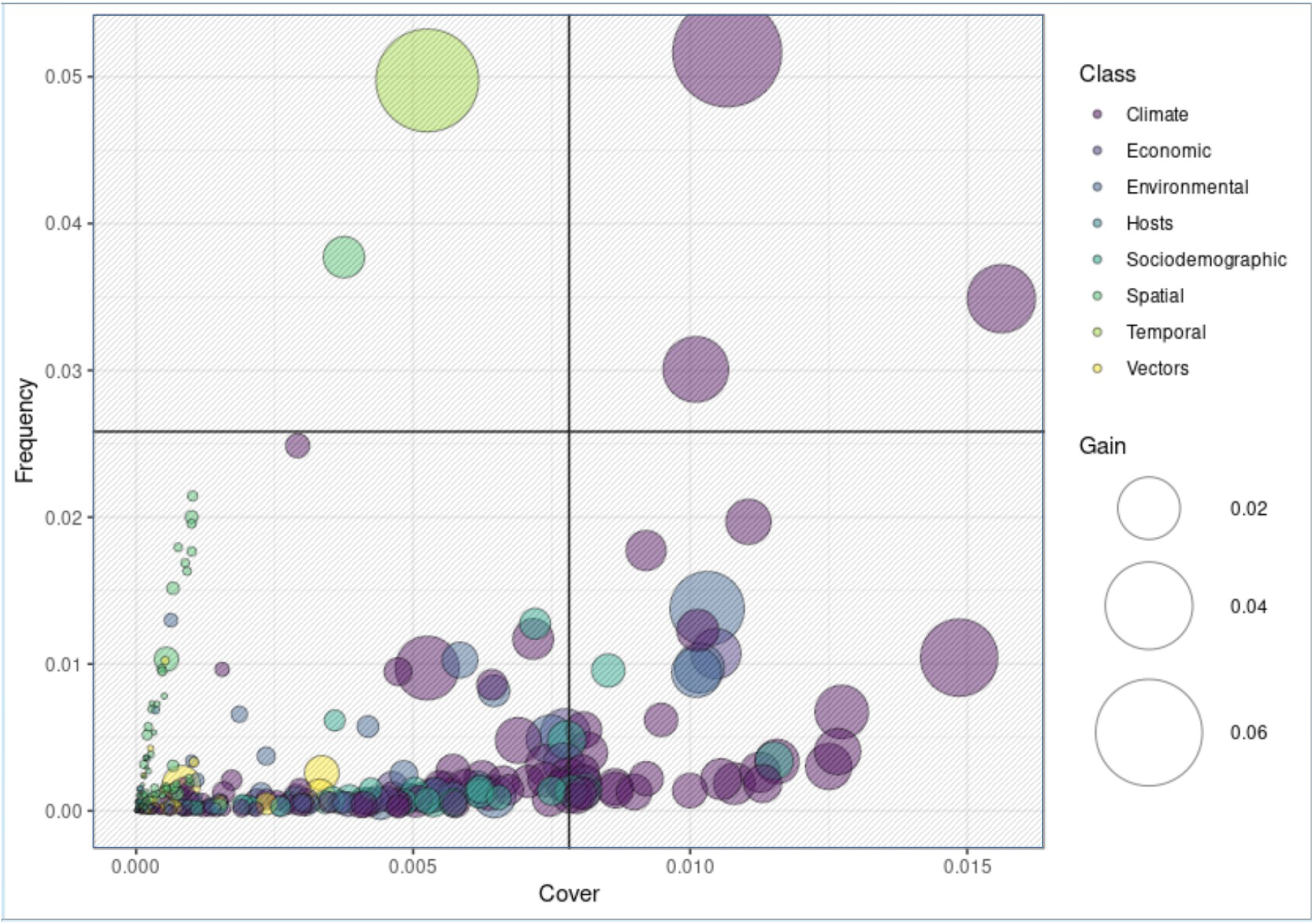
Imputed eature importance reflects research priorities. Here the feature and class dynamics underlying our XGB model are exposed. Each of three importance metrics are simultaneously plotted: “frequency” and “cover” along the y and x axis respectively and bubble size representing “gain”. Color represents feature class. Gain measures the overall impact of a feature as determined removing it and (re)evaluating model performance. Gain can therefore be thought of as an absolute effect estimate. As shown, most features impact a limited subset of regions (low cover). The features most likely to be relevant across cases are climatic (high cover). Models traditionally used for prediction generally cannot use variables that are not well represented in the data. Cover can therefore be thought of as a correlate to statistical significance. Frequency refers to the number of times a feature is found to add power to a prediction. A high frequency feature whose impact is not easily drowned out by other features. Features found in quadrant 1 are therefore not surprisingly those most likely to be found in published research studies: vapor pressure and maximum temperatures. Quadrant 2 features are all mostly covered in the literature, but not nearly as ubiquitously as those found in quadrant 1. These features represent overall lower proportions of total model variability, but are nevertheless mechanistically relevant. These second rank features imply a broad spectrum of climatic processes, including downward shortwave radiation in Q3 (3.1%), temperature seasonality (2.9%), vapor pressure in Q2 (2.4%), maximum temperature in Q3 (2.2%), and mean temperature of the warmest quarter (2.1%). Regional average economic wealth, measured in Euros and national currency equivalent, was found to be similarly important (1.3% each, 2.6% total). Land cover features commonly associated with mosquito breeding – industrial or commercial units (1.3%) as well as wet land habitats, such as water courses (1.2%) and inland marshes (1.2%) – rounded out the top. Quadrant 3 represents a substantial proportion of the features included this study. However, these low cover features arelikely to be determined “insignificant” in a traditional model and therefore prohibitively difficult to analyze. However, as shown, these features represent a substantial proportion of overall model output. Omission would therefore be expected to reduce and the predictive accuracy of any model. As shown, this variability can however be used by XGBoost’s ensemble processes. This ability to use such irregular sources of variability is arguably the source of XGBoost’s predictive power. However, adoption of XGBoost has been limited by the inability to generate true parameter estimates. The SHAP engine overcomes this limitation and therefore provides a means to generate statistically reliable effect estimates for even the poorest of samples.

The localized nature of the SHAP output allows for individual feature effects to be independently assessed for each case *(Figure 4a)*. This also allows for feature-wise model decomposition and assessment of aggregate effects specific to each feature class (*Figure 4b)*. Here, we confirm our previous observations regarding the aggregate effects of each feature class. Effects associated with climate are dominant and of about equal combined weight to all remaining features. Mapping furthermore confirms the differential and additives effect of each feature class with respect to geospatial outbreak risk contribution *(Figure S1)*. The mean magnitude of effect can furthermore be calculated for each individual feature, which is broadly equivalent to gain in the original model. We refer to this as the “SHAP score”. Based on this metric, the previously dominant autocorrelative feature was now removed to the fifteenth rank. Feature rankings otherwise remained broadly similar to those previously observed in the original XGBoost model *(Figure S2)*, therefore confirming that the SHAP reanalysis effectively preserves the global importance of each feature, while ensuring relative consistency of effect variability. At the local level, substantial bimodality of effect was noted for the top climatic features, which is consistent with the literature. Methods to mitigate the potential impact of such scale-dependent effects across regions are explored in subsequent sections.

**Figure 4.**
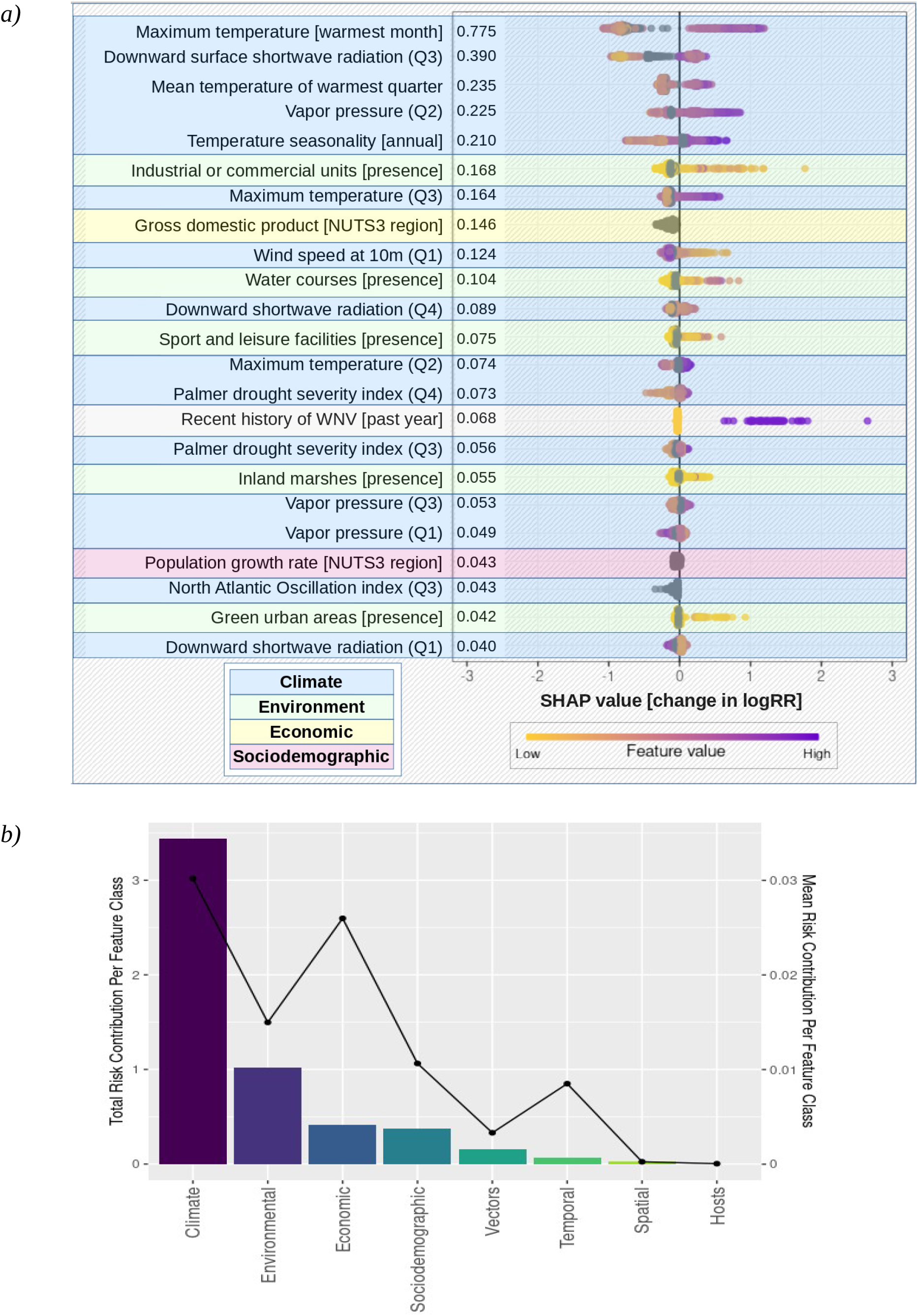
SHAP allows feature effect estimation across geospatial scales and user-defined contexts. (a) Case-wise effect predictions were generated for all features; here the the top ranked features are presented. This feature set represents the minimum number of features that captures 80% of output variability, as determined by a step-wise logistic regression process *(see Figure S4b)*. Notable bimodality of effect was observed in the highest ranking features, which were all climatic. This included the effects of maximum temperature of the warmest month, downward shortwave radiation in the third quarter, mean temperature of the warmest quarter, vapor pressure in the second quarter, and temperature seasonality. Bimodality is not unexpected given that the transformed surrogate data model simultaneously represents both indication and contraindication. (b) Feature class scores, including climatic variables, environmental variables, sociodemographic variables, economic variables, vector presence and diversity, host presence and diversity, spatial covariates, and temporal covariates (see *Supplementary Table 1*). The set of climatic features was found to contribute more to a positive prediction than all other feature classes combined (63%). Environmental features made up about half of the remaining risk pool (19%). The contribution of economic and sociodemographic features about equaled that of environment (8% and 7%, respectively; 15%, total). Vectors were overall less relevant at this scale, contributing about 3% of total risk. Even more so than vectors, the sum quantitative risk associated with spatial and temporal features (0% and 1%, respectively; 1%, total) and hosts (0%) were practically negligible at this resolution. Substantial differences were apparent in terms of mean absolute risk per feature class. The mean risk associated with climate and economic features were broadly similar (0.03 and 0.03, respectively). The risk contributions for sociodemographic features and temporal features, such as year and prior history of WNV infection, contributed far less on average (0.01 and 0.01, respectively). The average risk associated with vector, hosts, and spatial covariates were negligibly relevant at this scale (0.00, total).

### Case-wise stratification and hypothesis testing suggests mechanism driving 2018 outbreak

The statistical properties of the surrogate data model allow for segmentation and hypothesis testing, along with any other inferential analyses, including multivariate methods (see *Figure S2*). Here, this feature is exploited to conduct simple means testing *(Figure 6)* to determine which feature effects were stronger in the 2018 validation year compared to control (2010-2016). The feature strongly associated with increased outbreak risk in 2018 was vapor pressure (Q2), and it showed the largest relative increase by far in 2018. This coincided with a marked increase of the most commonly cited vectors, *Culex pipiens* and *Culex modestus*. In addition, the standardized effect of global climatic indices was also found to be substantially higher in 2018. This includes most notably, the North Atlantic Oscillation (NAO), variability of which has been associated with a broad range of infectious diseases in Europe. The protective effect of economic features, most particularly per person average income, was also found to be substantially attenuated in 2018 – considerably greater even than the effect size of climate.

**Figure 6.**
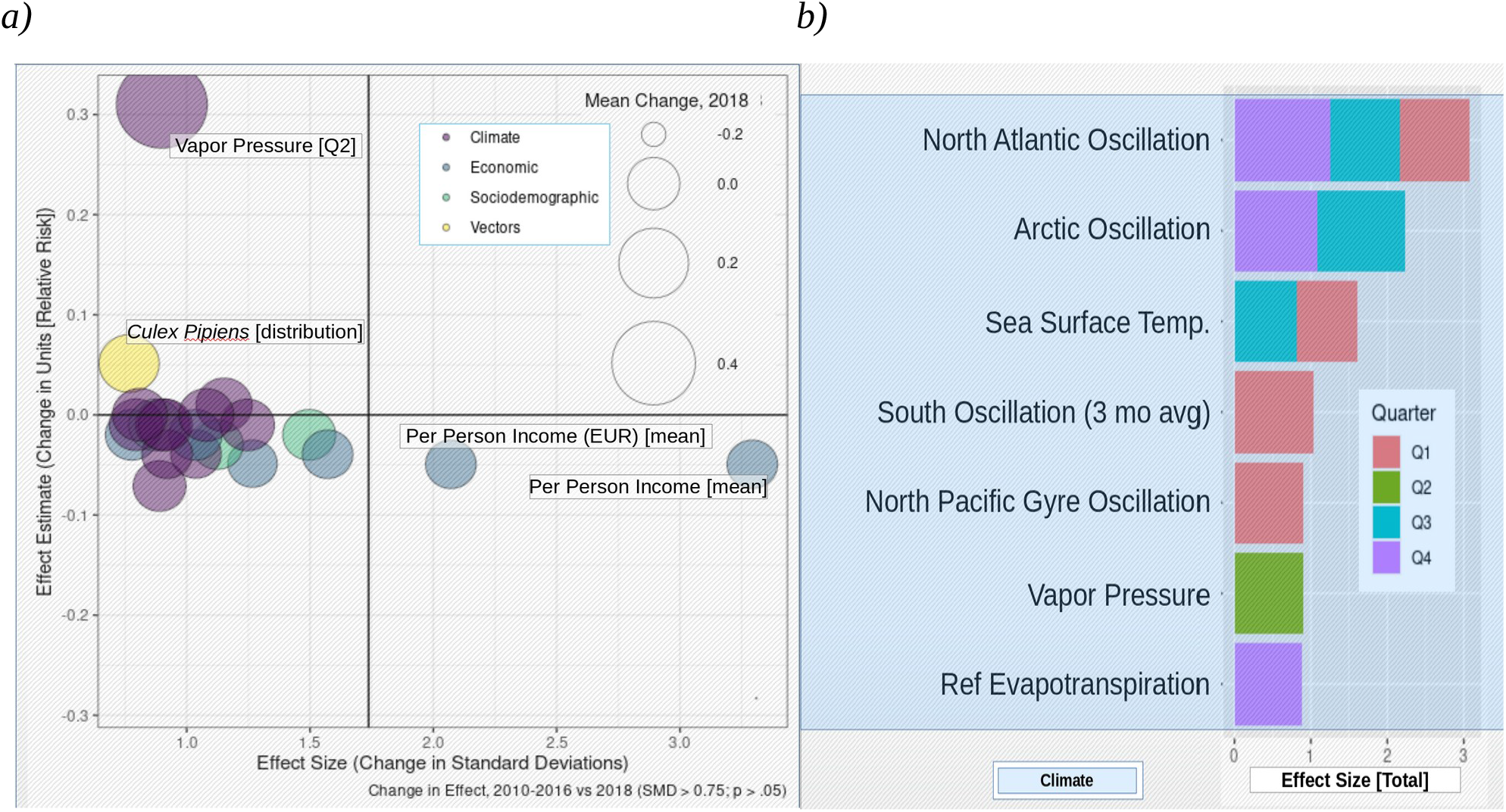
Statistical analysis of the SHAP effect matrix reveals drivers of extraordinary outbreak. (a) The effect of vapor pressure in Q2 of 2018 was found to be about twice that of the control period. Vapor pressure is commonly cited as the most indicative climatic feature with respect to local vectorial capacity and here too we find that the effect of the two most commonly cited vectors, *Culex pipiens and Culex modestus*, has also substantially increased at the same time. Wind speed in Q3, which has been associated with vector activity and distribution, was also found to contribute more substantially to outbreak risk in 2018. An increase in the effect of minimum and maximum temperature was noted for Q2, the time period cited most commonly as being critical with respect to life-cycle processes for mosquitos. The effect of drought, which has been associated with increased host-vector transmission due to increased contact rates between species due limited water resources, was also found to be attenuated Q3 through Q4. (b) Interestingly, the climatic features that showed the largest change in effect *size* (per standardized mean difference, SMD; refer to *Methods)* were the intradecadal climatic indices: the North Atlantic Oscillation (NAO, in Q1, Q3, and Q4) and Arctic Oscillation (AO, in Q3 and Q4). Both have been associated with increased outbreak risk in Europe, suggesting that regular variability in climate might possibly be a key underlying driver of apparently sporadic outbreaks in Europe.

**Figure 7.**
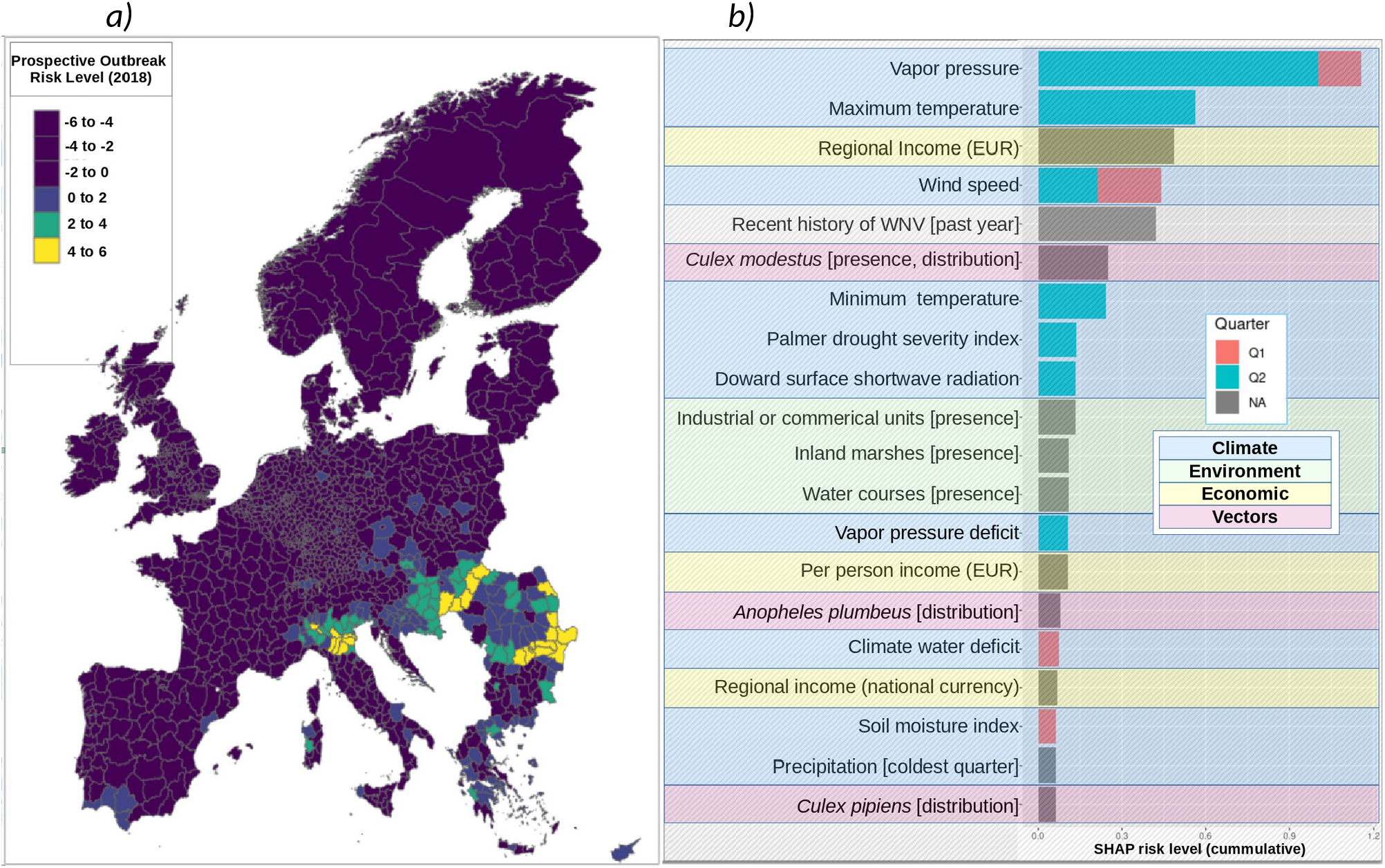
Prospective early warning model is similarly accurate, despite limited feature space. A model for prospective prediction was generated by omitting the feature specific to quarters one and two. This included the time-varying bioclimatic variables. Despite the limited feature space, model performance remained statistically identical to the explanatory model (AUC of 96%, sensitivity of 87% and balanced accuracy of 91%). Geospatial effect distribution is shown in *Figure 7a* and is broadly consistent with the exploratory model (see *Figure 2b*). Feature effect ranking, magnitude, and direction remained broadly consistent with the explanatory model, both overall and when feature effects are estimated for only those where outbreak was positively indicated (*Figure 7b*).

### Multivariate analysis suggests broad potential for expansion of WNV

A deterministic clustering procedure was applied the to stratify regions according to risk-based homogeneities (see *Methods)*. We refer to these clusters as “epidemiologic risk zones” (ERZ). Seven deterministically independent ERZ were identified, four major and three idiosyncratic *(Figure S7)*. Outbreak risk is strongly contraindicated in ERZ1 and ERZ2 due mostly to climate. Whereas ERZ5 corresponds with the region in which WNV is presently endemic. The present analysis therefore concludes by focusing on the ERZ most vulnerable to climate-driven changes in WNV outbreak risk, ERZ4. Findings suggest a region that is homogeneous with respect to increased outbreak risk driven by climate, environment, and host presence. Outbreak is presently contraindicated only due to insufficient presence of competent vectors (see *Discussion)*, an assessment with potential to suddenly change given the underlying risk structure demonstrated in this analysis.

### Prospective early warning model is similarly predictive and insightful

The explanatory model presented thus far has been used to identify associations and patterns indicative of increased WNV outbreak risk. However, it is not suitable for same-year prediction of outbreak risk, i.e. early warning. A derivative model was therefore constructed that included only those features measurable prior to the start of the WNV outbreak season. Despite the limited feature space, model performance remained statistically identical to the explanatory model (AUC of 96%, sensitivity of 87% and balanced accuracy of 91%; see *Figure 2a)*. Geospatial effect distribution *(Figure 7a)* indicated several regions beyond those typically indicated for WNV (see *Figure 1b)* and those indicated by the exploratory model (see *Figure 2b*). This is consistent with the slightly lowered sensitivity but may also reflect unreported cases. Feature effect ranking, magnitude, and direction remained broadly consistent with the explanatory model. This was also true when considering aggregate effects in only the regions where outbreak was positively indicated (*Figure 7b*). Here, distribution of *Anopheles plumbeus* was once again found to be a associated with increased WNV outbreak risk.

## Discussion

Several hypotheses have been proposed to explain the extraordinary expansion and severity of the 2018 WNV outbreak in Europe. One author concluded that aberrant temperature and drought preceded by anomalous precipitation in Q2 (i.e., a “wet spring”) were likely to be the leading cause^8^. Others have drawn similar conclusions, but the precise seasonal timeline varies region-to-region^41^. The present work lends support to such hypotheses. Indeed, even basic descriptives confirms the anomalous climate hypothesis *(Figure S3a)*. For example, we found that minimum and maximum temperature were significantly higher in 2018, for both Q2 and Q3 and vapor pressure reflected this same trend. We also found accumulated precipitation to be significantly lower in 2018 for both quarters. Distribution of the most commonly cited mosquito vector for WNV, *culex pipiens*, was also found to be higher in 2018. And indeed, this corresponded with a transmission season that was longer and more flatly distributed *(Figure S3b)*. The present work however expands upon this, by precisely confirming geospatial variability in determinant timing, while also robustly quantifying the overarching effect of large-scale climatic drivers and downstream causal dynamics.

### Overcoming the modifiable area unit problem

The “modifiable area unit problem” (MAUP) refers to the geospatial manifestation of a very basic statistical reality – observed means change in relation to the underlying distribution. And it has been noted now for well over 40 years as major confounder in geospatial analysis^25^. The present work, in addition to demonstrating a novel method to mitigate this effect, also demonstrates the degree to which the observed magnitude, ranking, and direction of effect changes as a function of geospatial scale and context. This was accomplished by exploiting a recently introduced method for reanalyzing a classification tree model to statistically homogenized effect matrix for use as a surrogate data model. As noted previously, the effect matrix is dimensionally identical to the original data but homogenized with respect to units, scale, and interpretation. Diagnostic analyses furthermore confirmed the surrogate data model to be statistically comparable with the original data (*Figure S4a*) but more robust with respect to tertiary multivariate analyses *(Figure S4b)*, particularly for smaller feature set sizes. The ability to robustly standardize for and compare the effect of scale-dependent variabilty is a direct product of these features. Such analyses are shown, not only to confirm associations previously only suggested in the literature, but also to allow for the discovery of novel associations at both global and locale-specific scales. This feature allowed for expert identification and quantification of the overarching determinative effect of large-scale climatic variability *(Figure 6b)*. This method furthermore allowed for the identification of several under-reported or plausible locale-specific host and vector populations *(Figure S5)*. Previous work similarly employed classification tree models to predict potential reservoir hosts for WNV^26^, but ours does so based on a direct assessment of contribution to disease outbreak risk. Taken together, this confirms the work of Cohen J.M. et al (2016)^27^ and others, who suggest that failure to address the MAUP may result in the effect of biodiversity and environment being downplayed in favor of climate.

### Assessing the potential impact of climate change on outbreak risk

The geospatial distribution of outbreak risk was shown to vary as a function of the feature class being observed. Our results confirm the primacy of climate as a mechanistic determinant of WNV outbreak risk. However, we also identified an equally influential constellation of features with a markedly different geospatial profile. The effect of non-climatic drivers, taken as a whole, extend northward well past the presently recognized limits of the zone commonly considered to be at risk for WNV. The risk-zone associated with non-climatic drivers extends well into the Baltics and parts of Scandinavia. Given the context of anthropogenic climate change, this is a finding that should not be discounted. These regions have historically been home to the similarly arthropod-borne illness malaria^28^. Furthermore, these latitudes are host to several species of trans-Saharan migrants from whom WNV has already been isolated^29^. Germany experienced its first autochthonous WNV cases in the year immediately following the first detection of WNV in local bird populations^30^. Indeed, according to our model for 2018, the regions in Germany affected by WNV in 2019 share a similar risk profile with the at-risk Baltic and Scandinavian regions. These results therefore suggest the potential for sudden changes in the geospatial range of WNV based on the preexistence of underlying non-climatic determinants. This work furthermore identified patterns of causation suggestive of the pivotal role of increased vectorial capacity due to regular climate variability. Most particularly, we found that a vector noted by the ECDC as being transmission capable but of low competence^31^ may have contributed substantially to the increased spread of WNV noted in 2018 (*Figure 5, S5)*. In addition, the present study also identified a trans-Saharan migrant whose range has shown notable increase over recent years *(Table S1b)*, likely due to anthropogenic change. These findings suggest that the true threat of changing climate may potentially lie in the “activation” of: otherwise dormant components of the transmission chain. Consequently, as seen in 2018, large-scale expansion of outbreak risk may occur far more suddenly than suggested by present climate change impact models^32-35^. Indeed, our clustering analysis revealed a sizeable area (ERZ4, see *Figure 7*) vulnerable to this very outcome. About one-tenth of the regions within this zone were affected by WNV in 2018. A diagnostic assessment revealed which factors were most strongly associated with this distinction, at this scale (see *Figure S6)*. Overall, as shown in *Figure 7c*, the underlying determinant structure suggests non-climatic factors presently contraindicate sustained transmission within this zone. Whether this might change due to shifts in vector populations due to anthropogenic changes or importation events is not immediately apparent. However, the similarity between ERZ4 and the historically endemic ERZ5 with respect to climatic risk suggest an urgent need to establish surveillance to prevent the establishment of more competent vector populations.

### Limitations

The scope of the present paper explicitly excludes elaboration as to the mathematical properties and interpretative consequences of the underlying estimation method used to produce these results. Lundberg, et al^19^ provide extensive methodological discussion on the present application; and the underlying theory, both with respect to local feature estimation and interpretation, has been established in literature and practice for well over 60 years^36,37^. Nevertheless, given the novelty and wide-ranging implications of the present application with respect to geospatial disease modeling, efforts to quantify the scope of interpretative application are considered to be a necessary next step.

## Conclusions

Recent years have seen substantial increases in the number of locales reporting West Nile virus (WNV) in Europe. The emergence of new lineages or strains has been another factor contributing to the increased number and severity of cases. Improved surveillance resolutions and predictive capabilities are therefore urgently needed. In this study we identify and describe the drivers of WNV outbreak risk in Europe using a novel method for generating fine-scale, local models. State of the art predictive performance and resolution are demonstrated using a novel methods in eXplainable AI. Novel analytical applications are demonstrated, including methods for: standardizing for scale-dependent feature effect variability, explanatory hypothesis testing, risk-based geospatial clustering, and even tertiary feature effect modeling. Results align well with the literature and several under-reported drivers are highlighted. Our results suggest anomalous variability in large scale climatic systems may have likely enhanced local climate suitability in 2018, partly driving increased spread and intensity of transmission in Europe.

The methods presented herein allow for an unprecedented degree of experimental efficiency. Given the parallelism among drivers of many infectious diseases, we expect broader scale adoption of such methods to lead to rapid advancements in capabilities to predict and control a broad array of outbreaks – which is crucial in light of changing climate and emerging infectious disease.

## Data Availability

All data is available upon request.

## Acknowledgments

A.A.G. received funding from the European Center for Disease Control and Prevention (contract no ECDC.9504). J.R. received funding from the Swedish Research Council Formas (grant nos. 2018-01754 and 2017-01300). We would like to thank the European Center for Disease Control for kindly providing funding, case and vector data, and support. And special thanks to the team at Exploratory.io who provided technical guidance and support.

## Author contributions

The authors worked collaboratively and contributed equally to the article.

---

**Figure S1.**
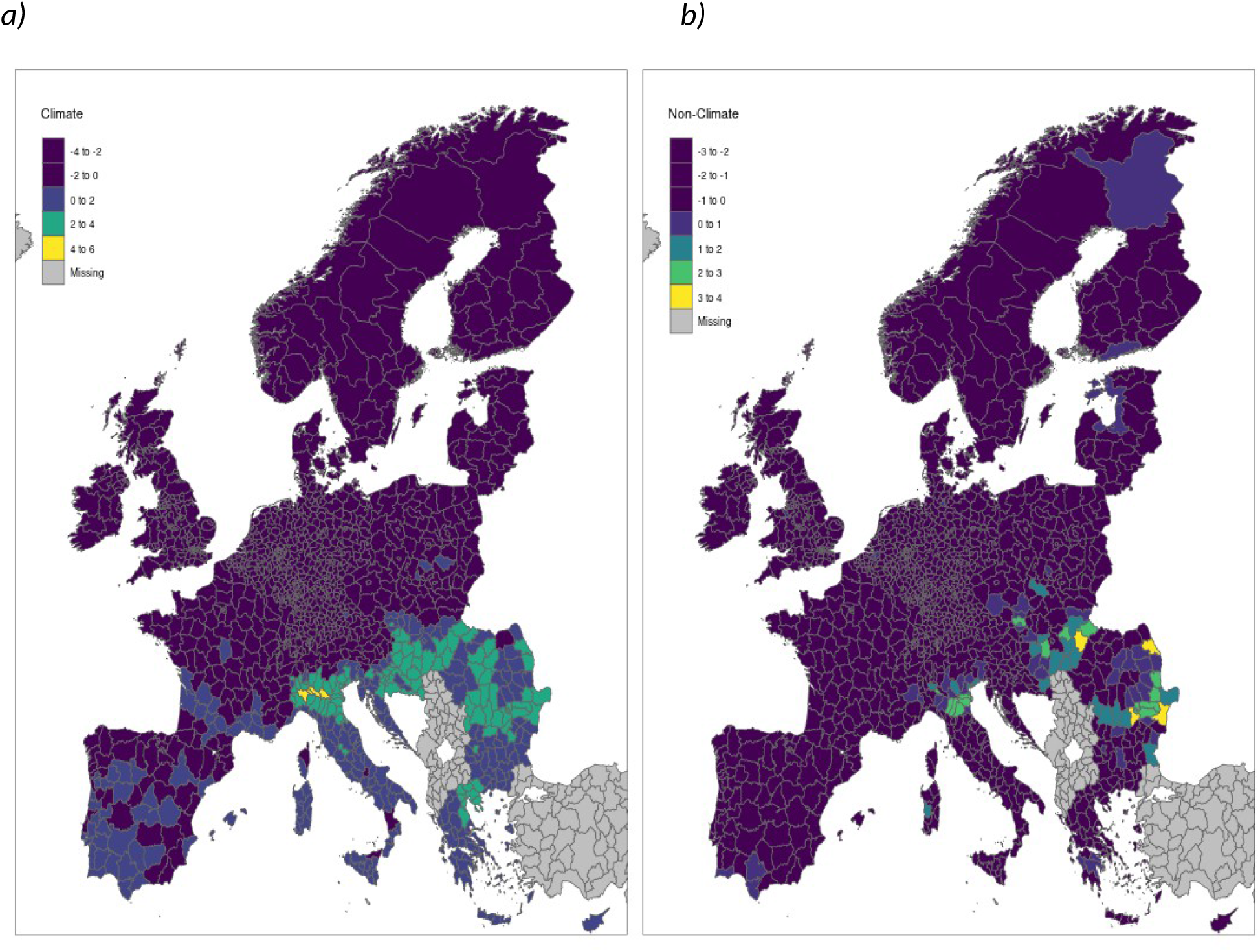
Model decomposition is a powerful application. The SHAP meta-model consists of an effect matrix that is dimensionally identical to the original, nominal value data set. The SHAP effect matrix can therefore be disaggregated on a case- or feature-wise basis and summarized accordingly. Here, feature-wise model decomposition reveals that the geospatial distribution of climatic and non-climatic features are markedly different. Non-climatic factors include economic, sociodemographic, vector, and host – associated features. Despite sum total aggregate effect being roughly equivalent to that of climatic risk, non-climatic risk extends into areas where WNV is otherwise presently contraindicated. This suggests that climate may, in some cases, serve to suppress non-climatic factors associated with increased outbreak risk. Were this to change, the emergence and spread of WNV and other similar diseases might occur far sooner than assumed under current climate change scenarios.

**Figure S2.**
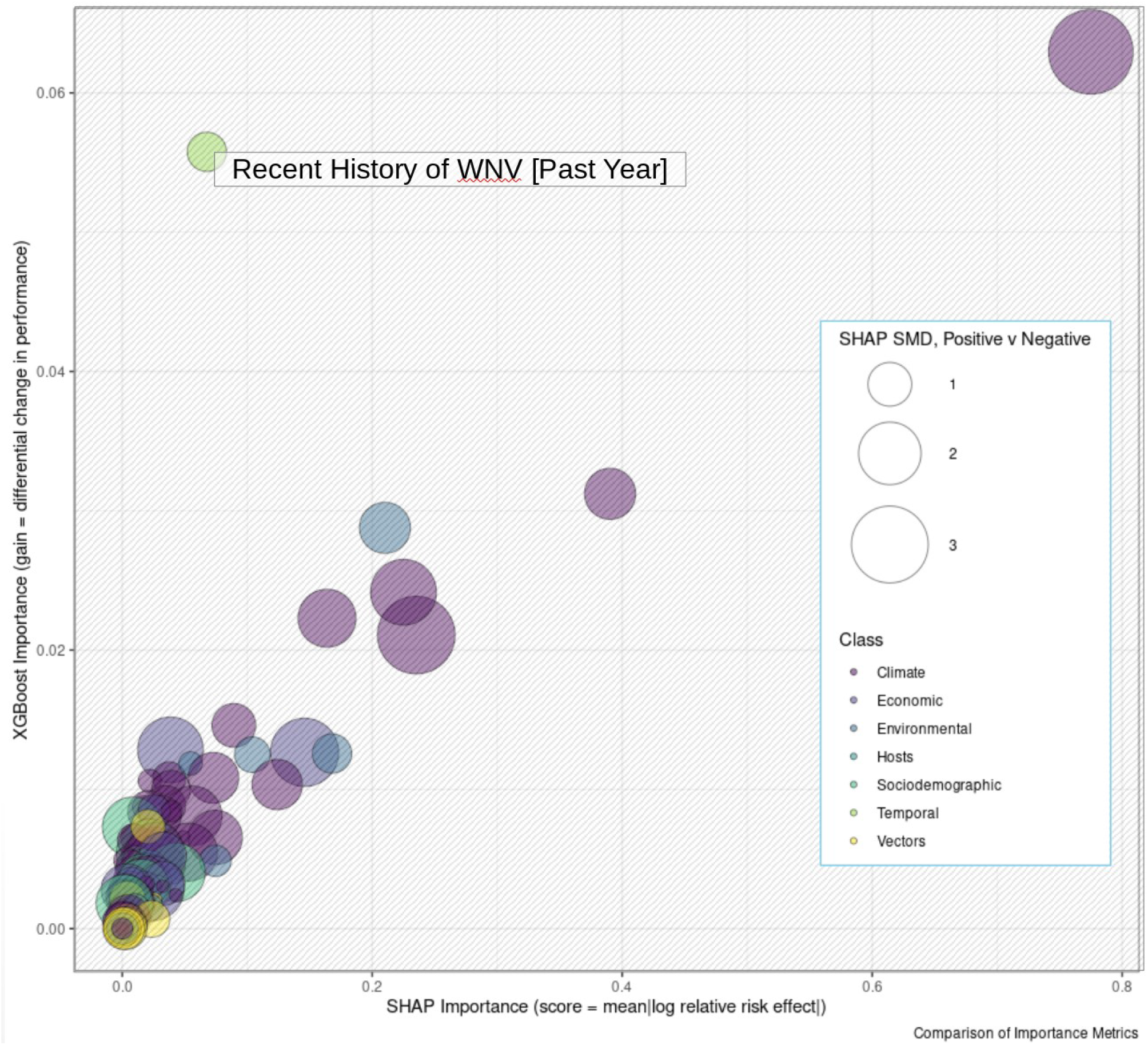
The SHAP reanalysis remains faithful to global model structure. In the context of classification tree models, “gain” refers to the degree to which global predictive performance is reduced is reduced when a given feature is removed from the model. Gain is one of the most common metrics used to evaluate feature importance in classification tree models. Feature importance can also be evaluated directly using SHAP by calculating the mean magnitude effect. Lundberg et al. (2020) refer to this as the SHAP score. Here, an almost one-to-one relationship is observed when gain (as calculated per XGBoost) is plotted against the SHAP score. The notable exception is the autocorrelative feature, “Recent History of WNV [Past Year]”. This feature was shown to be important only to a limited proportion of cases (*Figure 3*). This result demonstrates the degree to which the reannalysis of the XGBoost model substructure allows SHAP to modulate scale- or context-dependent variability of effect for features that are potentially of limited significance.

**Figure S3.**
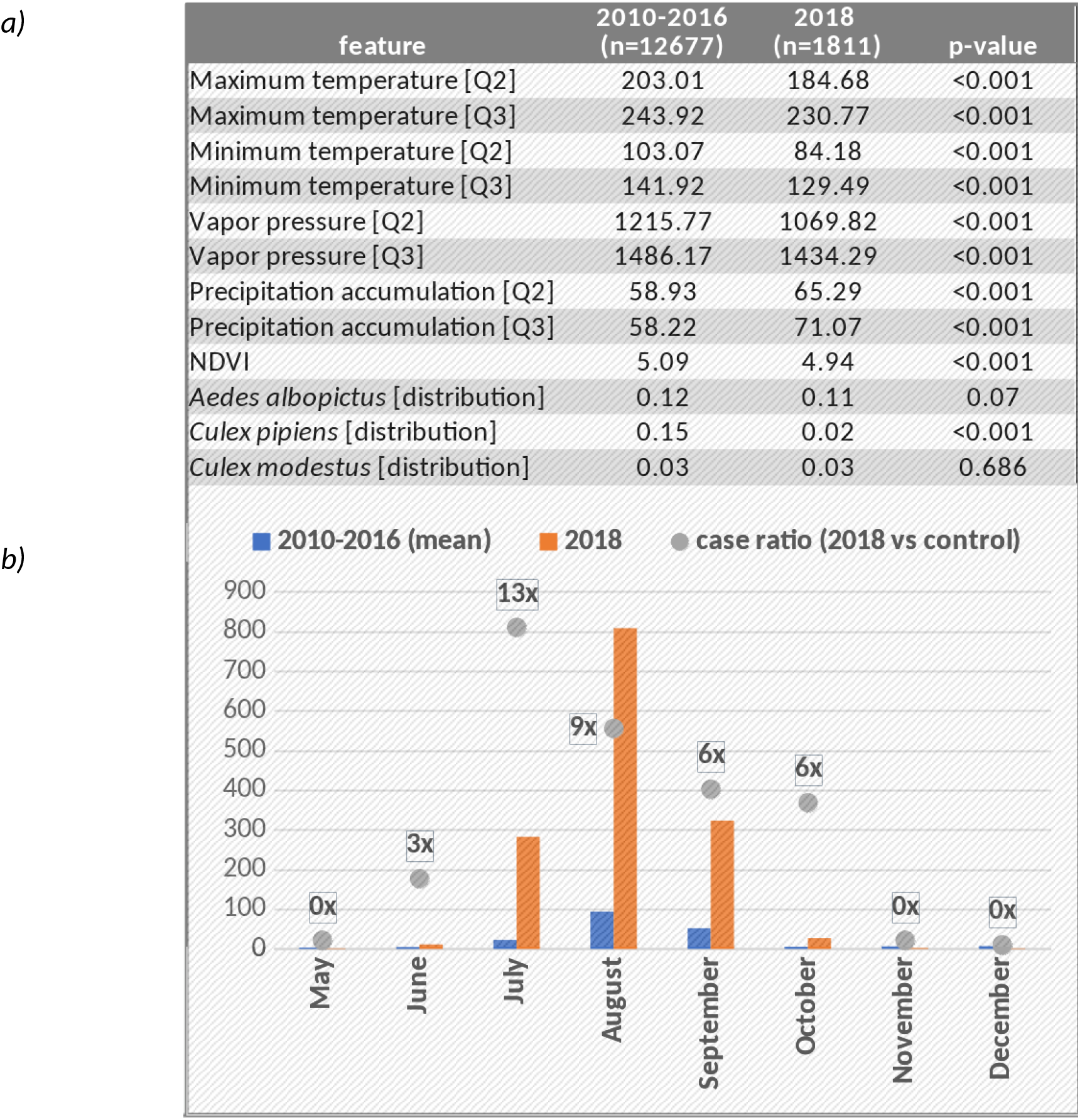
Means tests show the difference between the 2018 extraordinary event year in Europe and the 2010-2016 control period with respect to select feature values (*SF3a*). The differences reflect the consensus understanding of the factors coinciding with increased risk of vector-born diseases such as WNV. *SF3b* shows the extraordinary length of the 2018 outbreak season compared to the 2010-2016 control period in terms of absolute number of cases and the proportional increase in cases.

**Figure S4.**
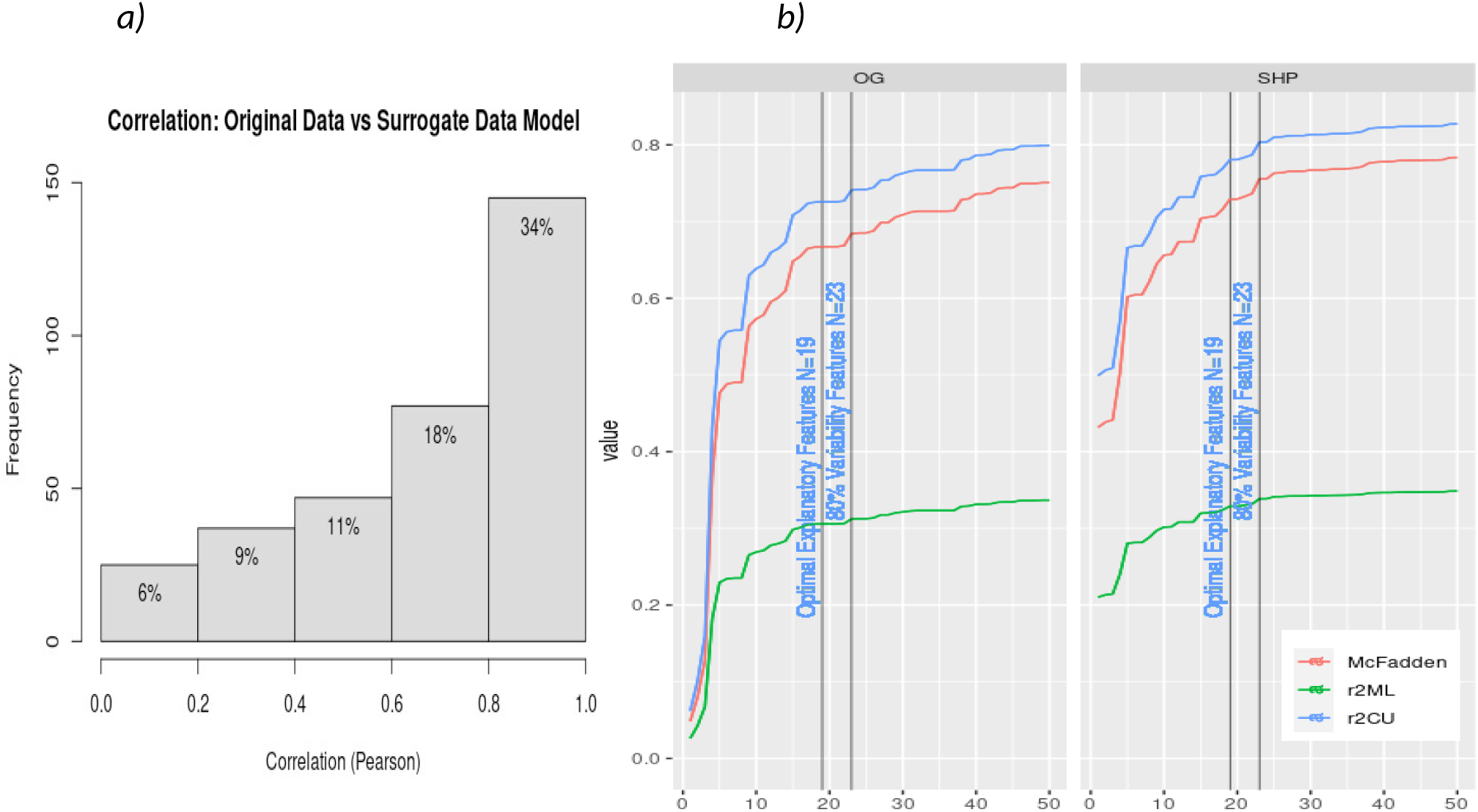
Surrogate data model aligns with but outperforms original data. The surrogate data model is heuristically designed to reproduce the output of the original (XGB) model while estimating the individual, localized effect of each feature. The surrogate data model is therefore dimensionally identical to the original data set but reflects deductively approximated effect estimates. We confirmed a high-degree of statistical correspondence between the two (*Figure 4a*). Stepwise logistic regression (*Figure 4b*) furthermore confirmed that the variability encoded within the surrogate data model aligned better with the original case data, for any given feature set size. This observed difference in predictive performance notably increases as model dimensionality is reduced. This finding is consistent across under various scenarios of model decomposition – e.g, by feature class.

**Figure S5.**
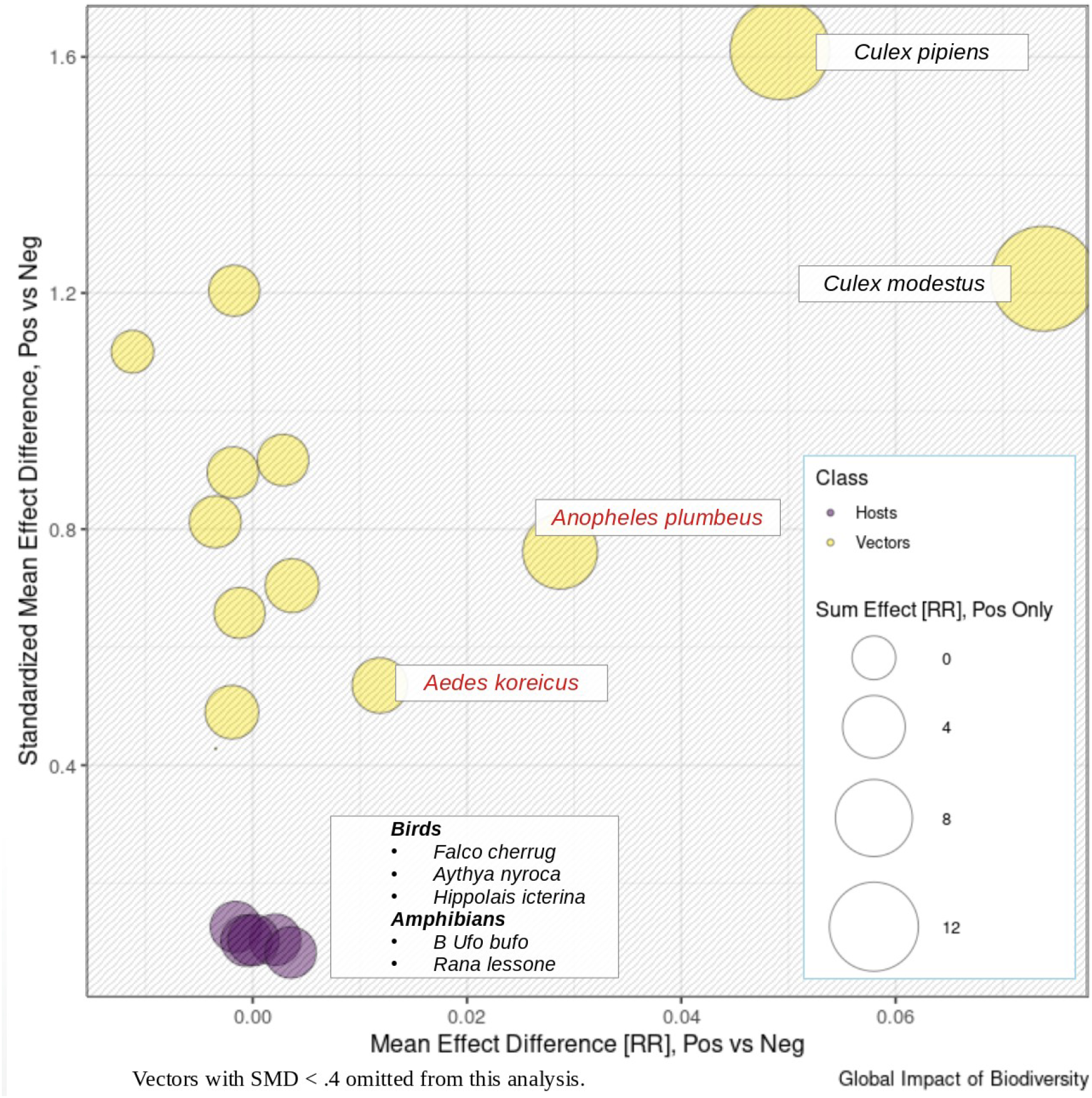
Broad diversity of hosts and vectors implied in Europe. Ranking and comparison of individual features with respect to aggregate effect is a simple matter using the surrogate data model. Means tests were conducted to comparing regions of positive and negative indication with respect to the effect of host and vector (p<.05). Effect sizes (standardized means difference) were calculated along with sum relatives risks for each, as an alternate measure of global effect. The likelihood of a comparatively complex European reservoir dynamic has long been suggested in the literature^8^, and again more recently^1^, and our results corroborate. The effect of vector distribution completely outstrips that of hosts. Which is suggestive of the primacy of anthropophilic mosquitos with respect to ultimate human transmission. The vectors most commonly associated with human transmission, *Culex modestus* and *Culex pipiens*, contribute more to aggregate risk than all other vectors combined. A notable runner-up is *Anopheles plumbeaus*, which has been cited by the ECDC as a potentially important vector. *Aedes koreicus* was also found to contribute marginally to outbreak risk. However, distinguishing it from its WNV-competent cousin, *Aedes japanicus*, has proven difficult and scant work exists to confirm its transmission status^55^. The remaining vectors have all been suggested by the literature, albeit with varying degrees of certainty^42,56,57^. For those exclusively involved in epizootic transmission, the effect on human outbreak risk is indirect and consequently of substantially lower impact. The effect of individual hosts was found to be minimal at the global scale. Most notable were the common toad *(Bufo buf)* and common pool frog *(Rana lessone)*, both of which have been found to contribute to in-the-wild infection of *Culex pipiens*, the primary WNV vector^58^. WNV has historically been isolated within a broad range of species and the possibility of enzootic transmission absent mosquito vectors has been confirmed^59^, which suggests the possibility of latent reservoirs awaiting the arrival of opportune climate and vectors.

**Figure S6.**
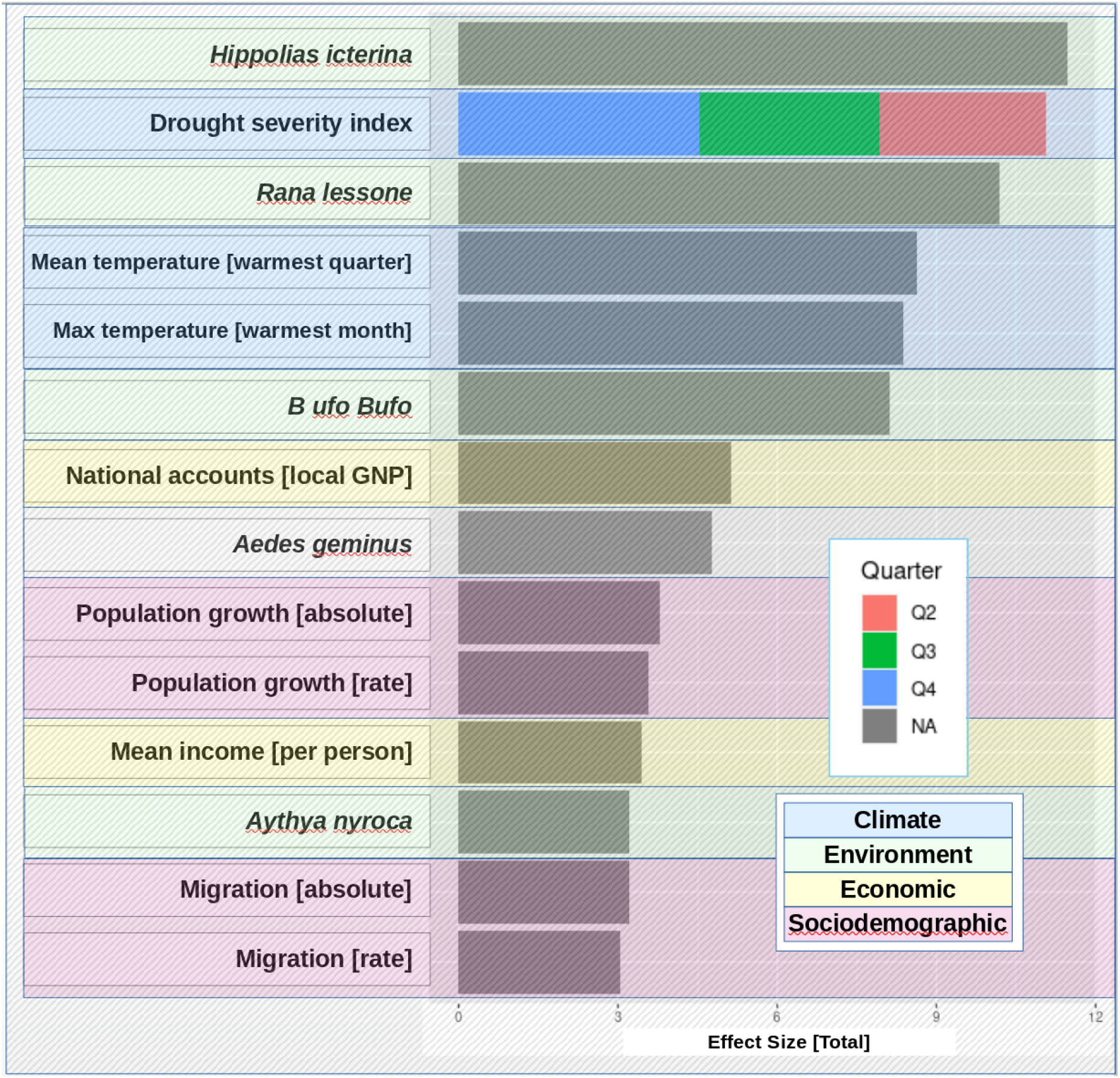
Host, economic, and sociodemographic effects are important at local scales. To understand the factors driving outbreak risk in ERZ4, scale-controlled effect sizes (standardized means difference, SMD) were calculated between regions of positive and negative indication. Effect sizes less than 3 (i.e., corresponding to an overlap limit of 7.2% and *r*^2^ of 0.69^62^) are omitted. Hosts, such as the trans-Saharan migrant warbler *(Hippolias icterina)*, the common pool frog *(Rana lessone)*, and the common toad *(Bufo buf)*, were found to be strongly associated with outbreak risk. Drought severity index, aggregated across quarters, was also found to be a major driver. Differences with respect to the effect of mean temperature of the warmest quarter and maximum temperature of the warmest month were similarly apparent. In addition, the effect of economic disparities, expressed in terms of region-level national accounts, were found to be substantial. Our local model furthermore identified increased risk associated *Aedes geminus*, a mosquito vector confirmed to feed on WNV-susceptible hosts including humans^60^, but otherwise generally not considered to be competent as a vector of WNV^61^. However, impact of this vector is only marginal, potentially indicating an indirect effect mediated through a role in zoonotic transmission of WNV.

**Figure S7.**
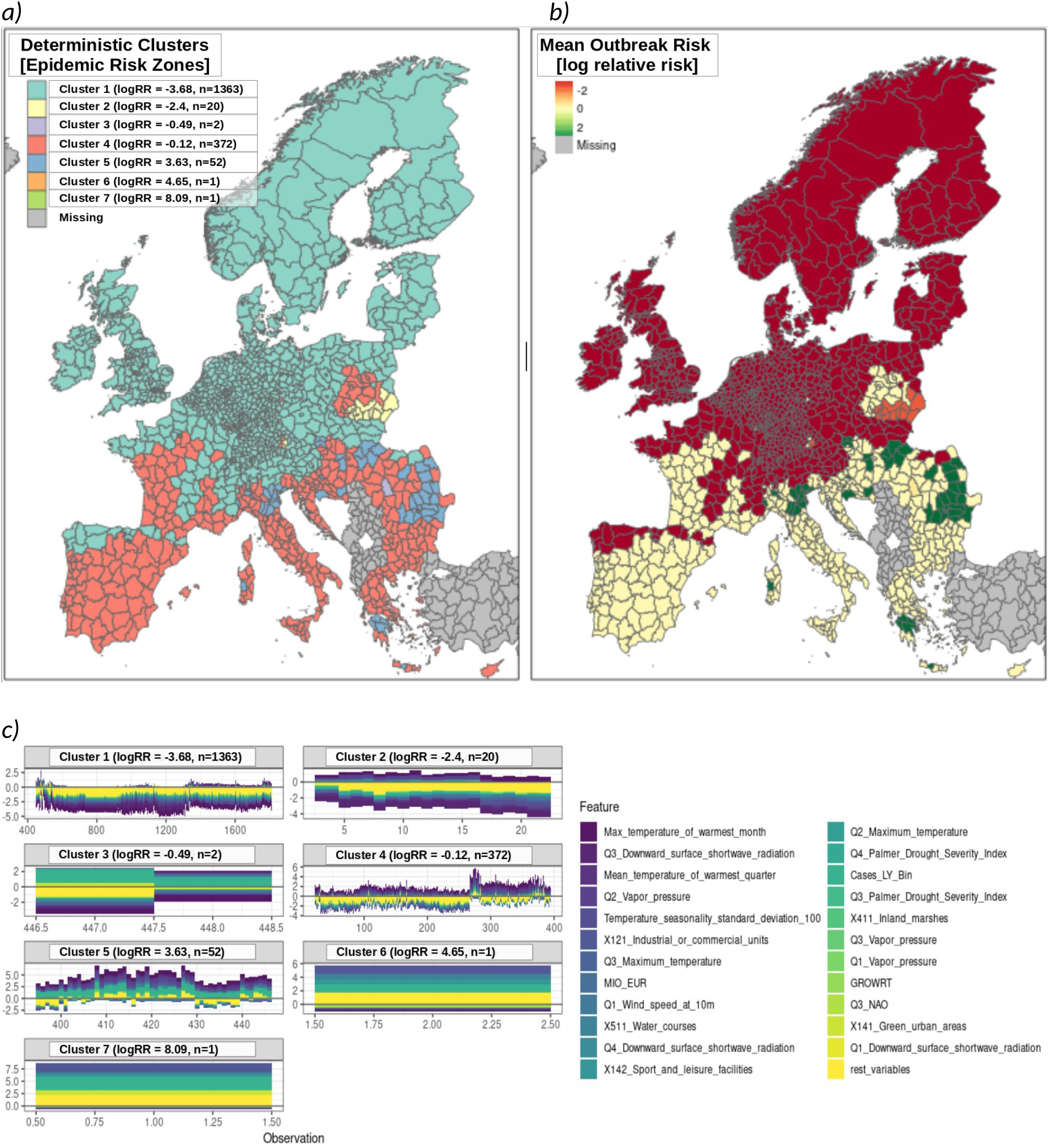
Effects predicted by Shapley transformation are mathematically robust. A clustering analysis revealed four major epidemic risk zones (ERZ) for WNV. Each risk zone was enumerated in ascending order according to mean relative risk (7a). ERZ1 consisted of the largest number of regions and largely coincided with the regions where outbreak was strongly contraindicated due to climate. ERZ2 consisted of a smaller, but structurally distinct, cluster of regions for which WNV is contraindicated. Significance testing revealed temperature seasonality and other climatic conditions to be consistent with increased risk of outbreak. However, this was found to be strongly contraindicated by apparently insufficient environmental, host, and vector related capacity. ERZ5 consisted of the historically endemic regions for WNV in Europe; these regions are largely uniform in terms of underlying epidemic risk structure. ERZ3, ERZ6, and ERZ7 were statistically unremarkable and are omitted from analysis. Risk-based mapping demonstrates the degree to which geospatial outbreak risk is indicated or contraindicated (*7b*). As indicated by its neutral risk profile, ERZ4 is climatically suitable for increased WNV outbreak risk. However, as shown in 7c, the underlying determinant structure suggests insufficient non-climatic risk to support sustained transmission within this zone. Whether this might change due to shifts in host or vector populations due to climatic or anthropogenic changes is not immediately apparent. However, the similarity between ERZ4 and historically endemic ERZ5 with respect to climatic risk suggest an urgent need to establish surveillance to prevent the establishment of more competent vector populations.

